# Risk Profile of Thanksgiving Gatherers and Subsequent SARS-CoV2 Testing and Diagnosis

**DOI:** 10.1101/2021.04.22.21255631

**Authors:** William You, Madhura Rane, Rebecca Zimba, Amanda Berry, Sarah Kulkarni, Drew Westmoreland, Angela Parcesepe, Mindy Chang, Andrew Maroko, Shivani Kochhar, Chloe Mirzayi, Christian Grov, Denis Nash

**Author notes:** **Corresponding Author:** William You, MSPH, Institute for Implementation Science in Population Health, City University of New York, 55 W 125th St, 6th Floor, New York, NY, 10027, United States.

## Abstract

**Background:** During Fall 2020 in the United States (U.S.), despite high COVID-19 case numbers and recommendations from public health officials not to travel and gather with individuals outside one’s household, millions of people gathered for Thanksgiving. The objective of this study was to understand if individuals’ behaviors and risk perceptions influenced their decision to gather, and if they did gather, their subsequent test seeking and diagnoses.

**Methods:** Participants were part of the CHASING COVID Cohort study - a U.S. national prospective cohort. The study sample consisted of participants who completed routine questionnaires before and after Thanksgiving. Non-pharmaceutical interventions (NPIs) use informed behavioral risk scores and a score of perceived risk of COVID-19 were assigned to each participant. Multinomial logistic regression models were used to assess the association between higher risk behaviors and gathering with other households, and the association of gathering with subsequent testing and test positivity.

**Results:** A total of 1,932 (40.5%) cohort participants spent Thanksgiving with individuals from at least one other household. Participants with higher behavioral risk scores had greater odds of gathering with one other household (aOR: 2.35, 95% CI: 2.0, 2.7), two other households (aOR: 4.54, 95% CI: 3.7, 5.6), and three or more other households (aOR: 5.44, 95% CI: 4.1, 7.2). Participants perceiving COVID-19 as a low-risk to themselves and others had greater odds of gathering with one other household (aOR: 1.12, 95% CI: 0.97, 1.3), two other households (aOR: 1.39, 95% CI: 1.1, 1.7), and three or more other households (aOR: 1.86, 95% CI: 1.4, 2.4). Those who spent Thanksgiving with one or more other households had 1.23 times greater odds (95% CI: 1.1, 1.4) of having a COVID-19 test afterward. There was no association between gathering for Thanksgiving and subsequent COVID-19 test positivity or developing COVID-19 symptoms.

**Conclusions:** Those who gathered with other households for Thanksgiving tended to engage in higher-risk activities. Thanksgiving gathering with other households was not associated with subsequently testing positive for COVID-19, but only a small proportion obtained post-travel testing. Public health messaging should emphasize behavior change strategies that promote safer gathering.

## INTRODUCTION

Pandemic control efforts in the United States (U.S.) were insufficient to prevent a second wave of the coronavirus disease 2019 (COVID-19) cases in late 2020. A combined approach of policy changes, public messaging, and non-pharmaceutical interventions (NPIs) were leveraged in response to increasing COVID-19 cases and deaths, but ultimately demonstrated limited effectiveness due to their lack of consistent use [1–5]. Public health officials repeatedly advised avoiding travel and gathering during the holiday season, and the U.S. Centers for Disease Control and Prevention (CDC) issued an advisory on November 19th, 2020, recommending that Americans should not travel for Thanksgiving [6], yet millions of Americans still traveled during the winter holiday season in November and December 2020 [7–9]. Data from the Transportation Security Administration (TSA) suggests air travel during the week of Thanksgiving was the highest since the beginning of the pandemic, about 10 times higher than the lowest rate of air travel in April, 2020 [9]. Ground travel increased almost 50% compared to earlier in the pandemic when shelter-in-place orders were implemented across the country [10,11].

Notably, in November and December 2020, when millions of people traveled and gathered for the holidays, the numbers of daily new COVID-19 cases in many states were among the highest reported to date, and U.S. case counts peaked in January 2021 [12]. Many have maintained that the second surge in COVID-19 daily new case numbers was largely caused by holiday travel and indoor gatherings among different households without proper protective measures [13,14]. While a few studies have shown holiday gatherings could have been associated with increased numbers of new COVID-19 infections [15], no published research to date has used longitudinal data to examine people’s decision-making around holiday gatherings and subsequent testing and diagnosis. The purpose of this analysis is to understand the relationships between individuals’ behaviors, risk perceptions, and decisions to gather with other households during Thanksgiving in late November 2020 and subsequent SARS-CoV-2 test seeking and diagnoses.

## METHODS

### Study design and participants

Study participants were screened and enrolled into the Communities, Households, and SARS-CoV-2 Epidemiology (CHASING) COVID Cohort study. The CHASING COVID Cohort study is a national prospective cohort study that was launched on March 28, 2020 to understand the spread and impact of the SARS-CoV-2 pandemic within households and communities. The survey methodology is described in detail elsewhere [16]. Study participants were recruited through social media platforms or through referrals using advertisements that were in both English and Spanish. The platform Qualtrics (Qualtrics, Provo, UT), an online survey platform widely used in social and behavioral research, was used for data collection.

The study conducted six rounds of routine data collection (Visit 1-Visit 6) in Project Year 1. The current sample consists of participants who completed a monthly follow-up survey (Visit 4) between November 17 and December 3, 2020, and a second monthly follow-up survey (Visit 5) between December 16 and December 28, 2020. The Institutional Review Board at the City University of New York (CUNY) approved the study protocol.

### Association between behavioral risk and risk perception and Thanksgiving gathering

In the first set of models, the outcome was Thanksgiving gathering, which was categorized into gathering with no other household, gathering with one other household, gathering with two other households, or gathering with three or more other households.

Behavioral risk-taking and COVID-19 risk perception were the main exposures of interest. The survey included a total of 42 questions related to the adoption of NPIs. We captured information on the use of protective gear (face masks, gloves, face shields, goggles), frequency of mask-use for indoor and outdoor activities, maintaining social distancing with known and unknown people, gathering in groups of 10 or more (attending protests, political rallies), spending time in public places (salons/gym/restaurants/places of worship), use of public transit, and air travel. The survey included 6 questions that captured participants’ perceptions about the risk of COVID-19 to themselves, their loved ones, and the community. Specifically, we asked participants if they stayed home from work when sick, if they self-quarantined, and the extent to which they were worried that they would get sick or reinfected with coronavirus, that their loved ones would get sick from coronavirus, and that coronavirus will overwhelm hospitals.

We assigned risk scores to each participant using their responses to the 42 behavioral risk questions and 6 risk perception questions in the Visit 4 survey (late November), as described above. For each question, we coded risk-affirmative responses as 1 and risk-averse responses as 0. Thus, higher scores reflected higher behavioral risks and lower perceived risk of COVID-19. For example, we asked participants how often they wore masks indoors during grocery shopping (indoor area with lots of people) in the past month. Those who responded with “Never” or “Sometimes” were coded as 1, and “Always” was coded as 0. We created behavioral risk and risk perception scores for each participant as the sum total of responses in each category. A participant could have a maximum behavioral risk score of 42 and a maximum risk perception score of 6. Median scores for each category were used to categorize participants’ higher/lower behavioral risk and higher/lower COVID-19 risk perception. Full details on the creation of risk scores can be found in the supplementary materials (sTable 1).

**Table 1:** Characteristics of CHASING COVID Cohort participants who gathered for Thanksgiving with other households.

|  | Thanksgiving with no other household (n=2,783) | %* | Thanksgiving with >=1 household (n=1,932) | % | Total (n=4,772)***** | % | p-values***** |
| --- | --- | --- | --- | --- | --- | --- | --- |
| **Median Age in years (IQR)** | 41 (32-56) |  | 37 (29-50) |  | 39 (31, 54) |  |  |
| **Gender** |  |  |  |  |  |  |  |
| Male | 1227 | 44.09 | 893 | 46.22 | 2138 | 44.80 | 0.2 |
| Female | 1473 | 52.93 | 989 | 51.19 | 2499 | 52.37 |  |
| Non-Binary | 83 | 2.98 | 50 | 2.59 | 135 | 2.83 |  |
| **Race/ethnicity** |  |  |  |  |  |  |  |
| NH White | 1714 | 61.59 | 1273 | 65.89 | 3000 | 62.87 | 0.04 |
| NH Black | 266 | 9.56 | 170 | 8.80 | 449 | 9.41 |  |
| Asian | 205 | 7.37 | 135 | 6.99 | 347 | 7.27 |  |
| Hispanic | 472 | 16.96 | 291 | 15.06 | 782 | 16.39 |  |
| Other | 116 | 4.17 | 56 | 2.90 | 176 | 3.69 |  |
| Missing | 10 | 0.36 | 7 | 0.36 | 18 | 0.38 |  |
| **Annual Household Income (in USD)** |  |  |  |  |  |  |  |
| Less than \$35,000 | 773 | 27.78 | 468 | 24.22 | 1265 | 26.51 | |
| \$35-49,999 | 308 | 11.07 | 204 | 10.56 | 518 | 10.85 | 0.05 |
| \$50-69,999 | 391 | 14.05 | 302 | 15.63 | 698 | 14.63 | |
| \$70-99,999 | 437 | 15.70 | 343 | 17.75 | 788 | 16.51 | |
| \$100,000+ | 791 | 28.42 | 566 | 29.30 | 1365 | 28.60 | |
| Missing | 82 | 2.95 | 47 | 2.43 | 135 | 2.83 |  |
| **Geographic region** |  |  |  |  |  |  |  |
| Northeast | 808 | 29.03 | 577 | 29.87 | 1399 | 29.32 | 0.006 |
| Midwest | 492 | 17.68 | 335 | 17.34 | 836 | 17.52 |  |
| South | 739 | 26.55 | 588 | 30.43 | 1345 | 28.19 |  |
| West | 714 | 25.66 | 418 | 21.64 | 1147 | 24.04 |  |
| US Territories | 3 | 0.11 | 2 | 0.10 | 5 | 0.10 |  |
| Missing | 27 | 0.97 | 12 | 0.62 | 40 | 0.84 |  |
| **Essential workers\*\*** |  |  |  |  |  |  |  |
| Yes | 369 | 13.26 | 256 | 13.25 | 631 | 13.22 |  |
| No | 2287 | 82.18 | 1609 | 83.28 | 3940 | 82.56 | 0.9 |
| Missing | 127 | 4.56 | 67 | 3.47 | 201 | 4.21 |  |
| **Median behavioral risk score at V4 (IQR)** | 10 (7-13) |  | 13 (10-17) |  | 12 (8-15) | <0.001 |  |
| **Median risk perception score at V4 (IQR)** | 1 (0-2) |  | 2 (0-3) |  | 1 (0-2) | <0.001 |  |

**Table 1: Characteristics of CHASING COVID Cohort participants who gathered for Thanksgiving with other households (cont'd)**
|  | Thanksgiving with no other household (n=2,783) | %* | Thanksgiving with >=1 household (n=1,932) | % | Total (n=4,772)***** | % | p-values***** |
| --- | --- | --- | --- | --- | --- | --- | --- |
| **Antibody positive at S1 (%)** |  |  |  |  |  |  |  |
| Reactive | 112 | 4.02 | 79 | 4.09 | 193 | 4.04 | 0.9 |
| Non-reactive | 1734 | 62.31 | 1207 | 62.47 | 2958 | 61.99 |  |
| Did not submit sample | 937 | 33.67 | 646 | 33.44 | 1621 | 33.97 |  |
| **Had symptoms at V4 (CSTE definition)\*\*\*** | 881 | 31.66 | 622 | 32.19 | 1520 | 31.85 | 0.7 |
| **Had symptoms at V5 (CSTE definition)** | 737 | 26.48 | 544 | 28.16 | 1291 | 27.05 | 0.2 |
| **Got tested at V4** |  |  |  |  |  |  |  |
| Yes | 584 | 20.98 | 497 | 25.72 | 1094 | 22.93 | <0.001 |
| No | 1965 | 70.61 | 1287 | 66.61 | 3281 | 68.76 |  |
| Tried | 136 | 4.89 | 71 | 3.67 | 211 | 4.42 |  |
| Missing | 98 | 3.52 | 77 | 3.99 | 186 | 3.90 |  |
| **Self-reported positive test prior to Thanksgiving\*\*\*\*** |  |  |  |  |  |  |  |
| Yes | 146 | 5.25 | 141 | 7.30 | 298 | 6.24 | 1 |
| No | 2637 | 94.75 | 1791 | 92.70 | 4474 | 93.76 |  |
| **Got tested at V5** |  |  |  |  |  |  |  |
| Yes | 660 | 23.72 | 566 | 29.30 | 1237 | 25.92 | <0.001 |
| No | 2006 | 72.08 | 1272 | 65.84 | 3310 | 69.36 |  |
| Tried | 95 | 3.41 | 77 | 3.99 | 178 | 3.73 |  |
| Missing | 22 | 0.79 | 17 | 0.88 | 47 | 0.98 |  |
| **Self-reported positive test at V5 among those who were tested\*\*\*\*\*** |  |  |  |  |  |  |  |
| Yes | 32 | 4.85 | 20 | 3.53 | 52 | 4.20 | 0.4 |
| No test | 491 | 74.39 | 428 | 75.62 | 926 | 74.86 |  |
| Waiting | 29 | 4.39 | 29 | 5.12 | 59 | 4.77 |  |
| Missing | 108 | 16.36 | 89 | 15.72 | 200 | 16.17 |  |
| **Thought they had COVID before Thanksgiving** | 462 | 16.60 | 386 | 19.98 | 855 | 17.92 | <0.001 |

\* col%
\*\* Essential workers include those who work in law enforcement, emergency management, retail (groceries, pharmacies, food), delivery, transportation, construction/agriculture, healthcare (including front office staff), school as teachers daycare, childcare
\*\*\* CSTE definition of CLI: At least 2 of these symptoms: fever, chills, rigors, myalgia, headache, sore throat, nausea, vomiting, diarrhea, fatigue, congestion, runny nose OR at least 1 of these symptoms: cough, shortness of breath, difficulty breathing, new loss of smell or taste
\*\*\*\* Reported a positive PCR test at V0, V1, V2, V3, or V4
\*\*\*\*\* Removed those who reported a positive test prior to V5 (n=28), i.e. counted only new incident cases
\*\*\*\*\* 57 responded with Do Not Know for the Question "Did you spend Thanksgiving with a person outside your household" and are not shown in this table
\*\*\*\*\* Chi squared p-values for categorical variables and Wilcox rank sum p-values for continuous variables
Abbreviations - NH: Non-Hispanic; IQR: Inter quartile range; V4: Visit 4 in November; V5: Visit 5 in December; S1: Sample collection 1 where dried blood spot samples were collected (May-September)

We also examined the association between Thanksgiving gathering and prior COVID-19 infection (if participants thought they had a COVID-19 infection at any time point between March 2020 and Thanksgiving), polymerase chain reaction (PCR) test at the prior visit (Visit 4), and COVID-19 symptoms at the prior visit (Visit 4).

### Association between Thanksgiving gathering and subsequent SARS-CoV-2 testing and diagnosis

Thanksgiving gathering in November was the main exposure and was dichotomized as gathered for Thanksgiving with no other household vs. with one or more households. The main outcome was self-reported SARS-CoV-2 PCR-test positivity (tested positive/tested negative/waiting for results) in the month prior among those who reported getting tested in the Visit 5 survey between December 16 and December 28, 2020. We only included incident cases reported at Visit 5 and excluded anyone who reported a positive PCR test in surveys prior to Visit 5.

Since only a quarter of participants reported getting tested post-Thanksgiving, we also examined if gathering with other households for Thanksgiving was associated with getting tested, as well as experiencing COVID-19 symptoms post-Thanksgiving.

### Statistical analysis

Chi-squared tests for categorical variables and Wilcoxon rank-sum test for continuous variables with corresponding *p*-values were used to describe the distribution of characteristics across participants who did and did not gather for Thanksgiving with other households. Multinomial logistic regression was used to estimate ORs and 95% CIs for all models. All models were adjusted for demographic factors such as age, gender, income, education, employment, and essential worker status. The model for the association between gathering for Thanksgiving and subsequent COVID-19 PCR positivity was additionally adjusted for participants’ behavioral risk score. Data were analyzed using R v4.0.1 [17].

## RESULTS

Of the 6,753 total CHASING COVID Cohort study members, 4,772 (70%) completed both the Visit 4 and Visit 5 surveys and were included in this analysis. 6 participants responded to the Visit 4 survey between November 26, Thanksgiving Day and December 3, 2020, one week later. Because the Visit 4 survey included questions about participants’ risk behaviors in the past month, which are relevant for this analysis, we included these 6 participants in the analysis. Participants with behavioral risk and risk perception scores greater than the median (12 for behavioral risk and 1 for risk perception) were categorized as “higher risk” or “lower risk perception”, respectively. Compared to other racial/ethnic groups, non-Hispanic White participants were more likely to be in the higher behavioral risk category (58.7% in low-risk and 68% in the higher-risk group; *p*-value <0.001). Lower income was associated with lower risk scores. In the lowest income category, 37.5% showed higher-risk behavior, compared to 50.1% in the highest income group; *p*-value <0.001)

In total, 2,783 participants (58.3%) reported that they did not spend Thanksgiving with other households, while 40.5% (n=1,932) reported gathering with at least one other household. Of these, 1,171 (60.6%) gathered with people from one other household, 484 (25.1%) with people from two other households, and 283 (14.6%) with people from three or more other households. The median behavioral risk score for those who did not gather with other households for Thanksgiving was lower compared to those who did (10 [IQR7-13] vs. 13 [IQR: 10-17]). The median risk perception score was also lower for those who did not gather with other households (1 [IQR 0-2] vs. 2 [IQR 0-3]) (Table 1). About one-third of the participants reported COVID-19 symptoms in the month prior to Thanksgiving, but there was no meaningful difference in the COVID-19 symptoms between those who gathered (32.2%) versus those who did not gather (31.7%). Of those who did not gather with other households, 21.0% reported receiving a test prior to Thanksgiving, compared to 25.7% among those who did gather with other households. A total of 324 of the 1,094 (29.6%) participants who tested prior to Thanksgiving responded that they were motivated to test prior to seeing friends or family.

Among the 1,923 participants who reported gathering during Thanksgiving, 141 (7.3%) had received a positive COVID-19 test between March 2020 and Thanksgiving; among the 2,783 participants who did not report gathering during Thanksgiving, 146 (5.3%) received a positive COVID-19 test in the same time period. In the post-Thanksgiving Visit 5 survey, only 25.9% of participants reported receiving a COVID-19 test, but testing was more common among those who gathered for Thanksgiving with members from other households (29.3% vs. 23.7%). Some participants reported a positive PCR test (n=52) and infection risk was lower among those who did versus did not gather with other households for Thanksgiving, but the difference was not statistically significant (3.5% vs. 4.9%, RR: 0.7, 95% CI: 0.4, 1.2). Of the 1,237 participants who reported a test in Visit 5, 59 (4.7%) got tested because they “attended gathering with more than 10 people”, of whom 4 (6.7%) tested positive; 139 (11.2%) got tested “after traveling”, of whom 4 (2.9%) tested positive; and 149 (12%) got tested “after seeing friends or family”, of whom 4 (2.7%) tested positive.

Participants categorized as having higher behavioral risk had a higher odds of gathering for Thanksgiving with one other household (aOR: 2.35, 95% CI: 2.0, 2.7), two other households (aOR: 4.54, 95% CI: 3.7, 5.6), and three or more other households (aOR: 5.44, 95% CI: 4.1, 7.2), compared to those categorized as lower behavioral risk, after adjusting for age, race/ethnicity, education, income, geographic area of residence, and essential worker status (Table 2, Model 1). Participants categorized as perceiving COVID-19 as a lower risk had a higher odds of gathering for Thanksgiving with one other household (aOR: 1.12, 95% CI: 0.97, 1.3), two other households (aOR: 1.39, 95% CI: 1.1, 1.7), and three or more other households (aOR: 1.86, 95% CI: 1.4, 2.4), compared to those perceiving COVID-19 as a higher risk, after adjusting for demographic variables (Table 2, Model 1). Those who thought they previously had COVID-19 (but were never diagnosed with a laboratory test) had higher odds of gathering with one other household (aOR: 1.33; 95% CI: 1.1, 1.6) as well as with three or more households (aOR: 1.56, 95% CI: 1.1, 2.2) compared to those who did not think they previously had COVID-19 (Table 2, Model 2). Having a negative test in the month prior to Thanksgiving and having COVID-19 symptoms during the period leading up to Thanksgiving were not associated with gathering with other households for Thanksgiving (Table 2, Models 3,4).

**Table 2:**
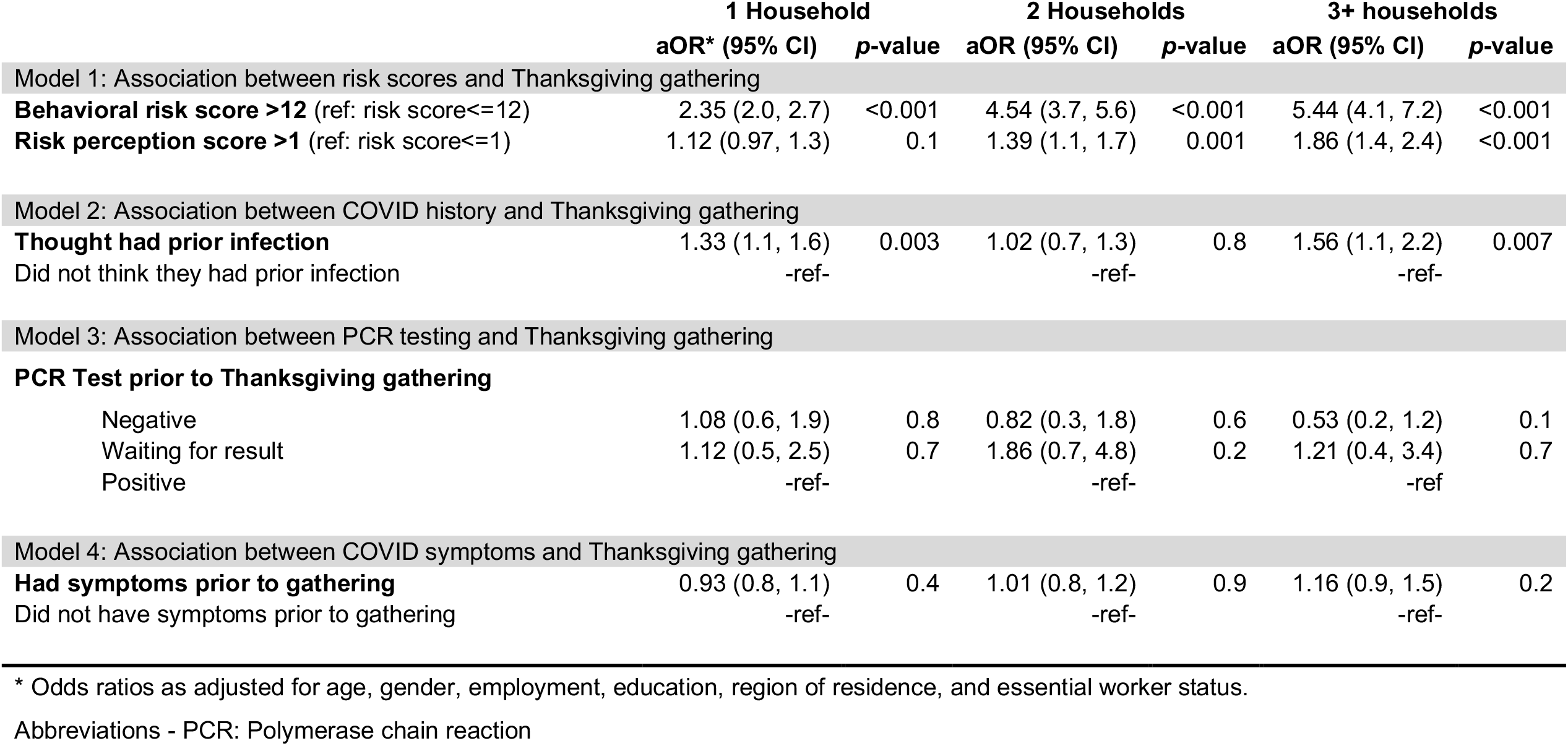
Results of multivariable models for association between risk scores, COVID history, and Thanksgiving gathering.

Those who gathered with one or more other households for Thanksgiving were more likely to get a COVID-19 test after gathering, compared to those that did not (aOR: 1.23, 95% CI: 1.1, 1.4) (Table 3, Model 1). However, the likelihood of testing does not differ by participants’ risk scores. Only 25.9% of the cohort received a COVID-19 test in Visit 5 post-Thanksgiving, of which 4.2% tested positive (Table 1). Post-thanksgiving testing was low, even among those who reported symptoms. Only 32.5% (420/1,291) of those who reported COVID-like symptoms post-Thanksgiving got tested. We did not find an association between gathering for Thanksgiving and subsequent self-reported COVID-19 infection or self-reported COVID-19 symptoms following the holiday among our participants (Table 3, Models 2,3).

**Table 3:**
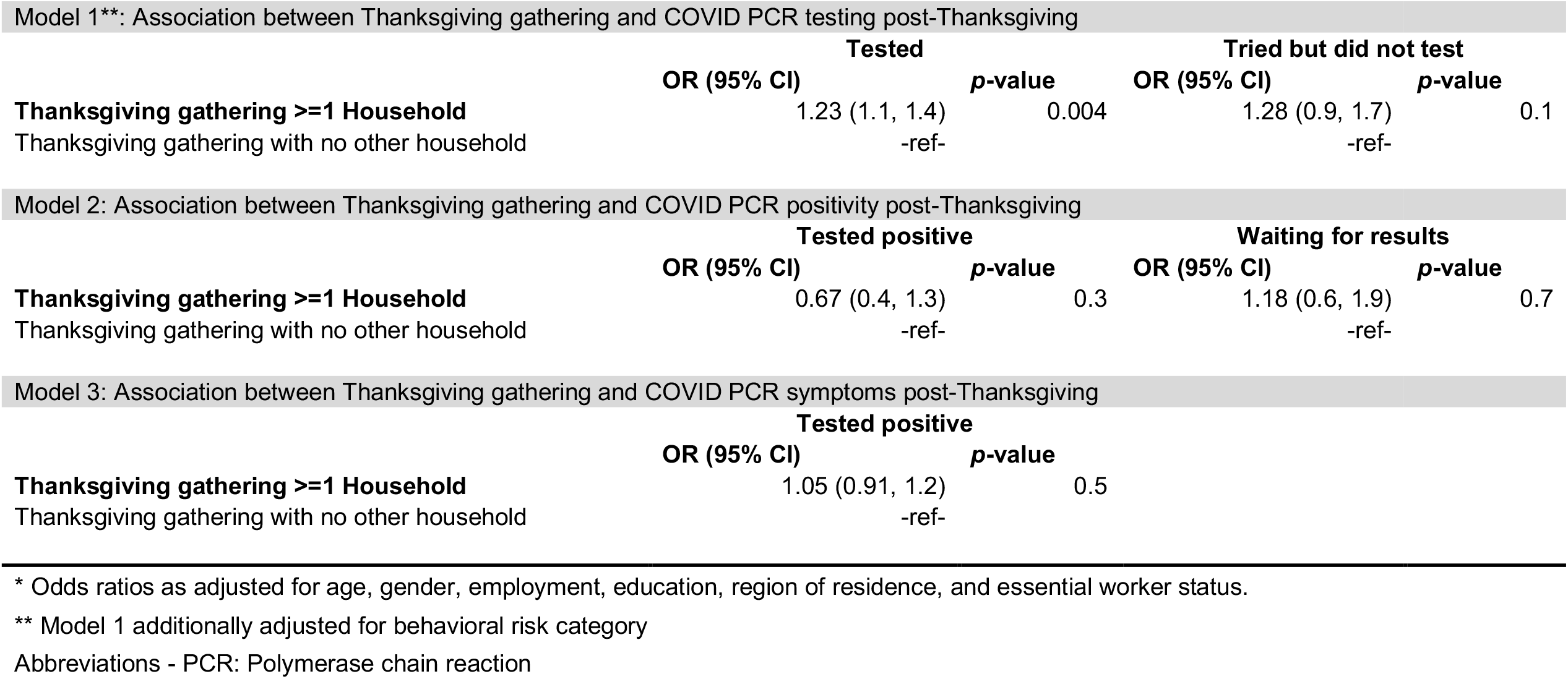
Multivariable models for association between Thanksgiving gathering with other households and PCR positivity for COVID.

Model 1\*\*: Association between Thanksgiving gathering and COVID PCR testing post-Thanksgiving
|  | Tested |  | Tried but did not test |  |
| --- | --- | --- | --- | --- |
|  | OR (95% CI) | p-value | OR (95% CI) | p-value |
| **Thanksgiving gathering >=1 Household** | 1.23 (1.1, 1.4) | 0.004 | 1.28 (0.9, 1.7) | 0.1 |
| Thanksgiving gathering with no other household | -ref- |  | -ref- |  |

Model 2: Association between Thanksgiving gathering and COVID PCR positivity post-Thanksgiving
|  | Tested positive |  | Waiting for results |  |
| --- | --- | --- | --- | --- |
|  | OR (95% CI) | p-value | OR (95% CI) | p-value |
| **Thanksgiving gathering >=1 Household** | 0.67 (0.4, 1.3) | 0.3 | 1.18 (0.6, 1.9) | 0.7 |
| Thanksgiving gathering with no other household | -ref- |  | -ref- |  |

Model 3: Association between Thanksgiving gathering and COVID PCR symptoms post-Thanksgiving
|  | Tested positive |  |
| --- | --- | --- |
|  | OR (95% CI) | p-value |
| **Thanksgiving gathering >=1 Household** | 1.05 (0.91, 1.2) | 0.5 |
| Thanksgiving gathering with no other household | -ref- |  |

\* Odds ratios as adjusted for age, gender, employment, education, region of residence, and essential worker status.
\*\* Model 1 additionally adjusted for behavioral risk category
Abbreviations - PCR: Polymerase chain reaction

## DISCUSSION

Travel and indoor gatherings with other households during the Thanksgiving holiday may have contributed to the already increasing numbers of COVID-19 cases and deaths in the U.S. [18]. This study explored different behavioral and risk perception factors and their associations with gathering during Thanksgiving in November 2020. Participants who perceived COVID-19 to be a lower risk to themselves and others and those who regularly engaged in higher COVID-19 risk activities the month prior to Thanksgiving were more likely to gather with people outside their households at Thanksgiving. Having COVID-like symptoms and test positivity prior to and post-Thanksgiving gathering were not significantly associated with gathering at Thanksgiving, but a substantial proportion (22.9%) of participants tested for SARS-CoV-2 prior to Thanksgiving, and the point-prevalence was not insubstantial (3.5%) among those who did gather and tested on their return.

Although lacking coordination and consistency, the public health message from various sources advising not to gather during Thanksgiving was strong. However, the CDC did not advise against Thanksgiving holiday travel until one week before Thanksgiving in 2020 [6]. By then, many people had already booked flights or made travel plans [19]. More timely and coordinated guidelines at the federal, state, and local levels were necessary during the holiday season when travel was anticipated, and could have included pre- and post-travel testing and quarantining, as the CDC currently recommends for unvaccinated persons [20].

Travel often involves passing through crowded places such as train stations or airports, and using public transportation where social distancing can be difficult to maintain. Although 91% of long-distance holiday travel occurs by personal vehicles, such as by car, according to the U.S. Department of Transportation, the average Thanksgiving long-distance trip length is 214 miles [21], seven times longer than the distance an average driver travels on a daily basis [22]. People with symptomatic and asymptomatic COVID-19 who travel and gather with other households have the potential to spread the virus beyond their local communities, which makes contact tracing even more challenging than usual. Regulations related to traveling, testing, and quarantine vary by jurisdiction, which can be confusing for travelers [23]. A set of simple, clear, and standard national-level COVID-19 prevention guidelines for interstate and long-distance travelers could be easier for travelers to adopt and practice consistently. State and local governments could choose to strengthen the established federal standard if additional preventive measures are deemed necessary, given local or other contextual considerations.

A quarter of study participants who gathered at Thanksgiving reported receiving a COVID-19 test in the month prior. Participants may have encountered barriers to testing such as higher than usual demand for COVID-19 tests before Thanksgiving, reducing availability, and increasing wait time [24]. It is concerning that the majority of the respondents who gathered at Thanksgiving did not know their COVID-19 status immediately prior to gathering, a third of whom also experienced COVID-related symptoms in the month before gathering. Depending on timing, some of the above-mentioned participants should have self-isolated if COVID-related symptoms began during the 14 days leading up to Thanksgiving. However, given the virus’s incubation period, negative COVID-19 test results should not serve as a green light for safe indoor gatherings without masks [25].

Notably, in this analysis, receiving negative test results prior to Thanksgiving and having COVID-19 symptoms during the period leading up to Thanksgiving were not associated with gathering with other households at Thanksgiving. Our data suggest that many in our cohort appeared to have used testing pre- and post-Thanksgiving as a strategy for risk reduction. 4.9% of those who stayed home for Thanksgiving tested positive for COVID-19 in the month prior, suggesting the result of the test may have driven some participants to change their plans and stay home. However, since there was no clear guidance on this from public health authorities, the use of this strategy may have been uninformed at worst or suboptimal at best, with neither leading to optimal risk reduction. Moreover, many who could have reduced risk through testing did not and visited with other households anyway. All of these scenarios should be anticipated by public health authorities, and directly addressed with/incorporated into pandemic-related public health messaging in the future. Therefore, in addition to encouraging the public not to gather, early and consistent messaging related to interventions around how to more safely gather during major holidays may have been more effective in preventing new infections. Strategies used in reducing COVID-19 transmission at indoor gatherings, such as mask use at home, open windows to increase air circulation, and reduced close contact can be transferable in the context of holiday gatherings with different households [26,27]. In addition, heightened sequestering or full quarantine in the 10-14 days leading up to such gatherings, when feasible, is a very effective strategy for safer gathering.

Although the self-reported PCR positivity after Thanksgiving among those who gathered for Thanksgiving was lower compared with those who did not gather in our cohort, a substantial proportion of those who gathered still tested positive after Thanksgiving (3.5%). If this proportion extends to the untested cohort, a considerable number of people could have been infectious while/after visiting other households during Thanksgiving. However, our ability to detect an association between positivity and gathering was limited by the small number of participants who reported receiving a COVID-19 test after Thanksgiving. In contrast to our results, a study conducted by Mehta et al. after Thanksgiving in 2020 found that those who celebrated Thanksgiving with non-household members or celebrated outside their home were significantly more likely to report COVID-19 symptoms, testing for SARS-CoV-2, and testing positive for SARS-CoV-2 infection, compared to those who had Thanksgiving with household members only [15]. In another study using serologic data from the CHASING COVID Cohort, we found an increased risk of seroconversion among participants who reported gathering with large groups indoors and those who traveled by air [27]. A lack of testing after such gatherings also makes it less possible for people to self-quarantine/isolate when exposures did occur. Therefore, pandemic mitigation efforts that focus on limiting large gatherings or emphasizing the importance of NPIs and testing are still needed. Even as the vaccine rollout continues, vaccinated individuals should continue to wear masks, maintain physical distance, and practice other prevention measures when visiting or gathering with unvaccinated vulnerable populations [28].

Several limitations should be considered when evaluating the results of this study. We used a self-administered survey and the data on both risk behaviors and COVID-19 infection status could be subject to self-report bias. Participants’ answers on gathering and testing behaviors may have also been impacted by social desirability bias. Additionally, only a limited number of participants reported getting tested before and after Thanksgiving. Hence, it is difficult to estimate the actual incidence of COVID-19 infection due to gathering with other households. However, by assessing other behaviors and COVID-19 symptoms, we were able to assess the potential risk of transmission. Finally, it is possible that risk-averse participants are more likely to get tested and test negative. Therefore, the positivity rate among testers after Thanksgiving gatherings could have been an underestimate of the true positivity rate among Thanksgiving gatherers in our study.

### CONCLUSIONS

## Data Availability

Data are available upon request.

Those who participated in Thanksgiving gatherings with other households tended to regularly engage in higher COVID-19 risk activities prior to Thanksgiving. For the current and future pandemics, a set of simple, clear, and standard national-level COVID-19 prevention guidelines for interstate and long-distance travelers to prevent spread, along with well-timed public health messaging that incorporates guidance on testing and quarantine (pre/post-travel), would likely help reduce risk at and following indoor gatherings. In addition to public health messaging, future research should also emphasize behavior change strategies that promote safer gathering via risk reduction.

