## Supplemental Tables for "Risk Profile of Thanksgiving Gatherers and Subsequent SARS-CoV2 Testing and Diagnosis"

**sTable 1. Scoring of COVID-related risk behavior and risk perception**

| Question in survey | Behavioral risk | Risk perception | Score |
|---|---|---|---|
| **Have you traveled by plane since March 2020?** | x | | 1 if Yes; 0 if No |
| **In the past month, how often did you wear a mask indoors during the following activities?** | | | |
| Grocery shopping (indoor area with lots of people) | x | | 1 if Never or Sometimes; 0 if Always |
| While visiting friends/family who are not part of your household | x | | 1 if Never or Sometimes; 0 if Always |
| While at work (inside an office building or other work environment) | x | | 1 if Never or Sometimes; 0 if Always |
| Using public transit (bus, train, subway) | x | | 1 if Never or Sometimes; 0 if Always |
| Visiting a salon, gym (an indoor area with a few people) | x | | 1 if Never or Sometimes; 0 if Always |
| **In the past month, how often did you wear a mask outdoors during the following activities?** | | | |
| Visiting friends/family outside of your household | x | | 1 if Never or Sometimes; 0 if Always |
| Exercising or walking on the street | x | | 1 if Never or Sometimes; 0 if Always |
| At an outdoor gathering, such as a sporting event, political rally, concerts | x | | 1 if Never or Sometimes; 0 if Always |
| **In the past month, have you done any of the following as a result of concerns about the new coronavirus? For each item select Yes, No, Or Not Applicable.** | | | |
| Avoided shaking hands or hugging | x | | 1 if No; 0 if Yes; Not Applicable was not included |
| Worn gloves | x | | 1 if No; 0 if Yes; Not Applicable was not included |
| Worn a face mask | x | | 1 if No; 0 if Yes; Not Applicable was not included |
| Worn a face shield | x | | 1 if No; 0 if Yes; Not Applicable was not included |
| Worn safety goggles | x | | 1 if No; 0 if Yes; Not Applicable was not included |
| Cleaned and disinfected frequently used objects or surfaces (for example a smartphone) | x | | 1 if No; 0 if Yes; Not Applicable was not included |
| Avoided touching your face | x | | 1 if No; 0 if Yes; Not Applicable was not included |
| Avoided public transportation | x | | 1 if No; 0 if Yes; Not Applicable was not included |
| Physically separated from people within your household (renting a separate home or staying on a separate floor or room) | x | | 1 if No; 0 if Yes; Not Applicable was not included |
| Avoided public transportation | x | | 1 if No; 0 if Yes; Not Applicable was not included |
| Avoided gatherings with people outside your household | x | | 1 if No; 0 if Yes; Not Applicable was not included |
| Formed a pod or a team (a group of people who all agree to only socialize with each other) | x | | 1 if No; 0 if Yes; Not Applicable was not included |
| Made plans to protect older persons that you know (arranged delivery of food or medicine) | x | | 1 if No; 0 if Yes; Not Applicable was not included |
| Stayed home from work when you were sick | | x | 1 if No; 0 if Yes; Not Applicable was not included |
| Self-quarantined | | x | 1 if No; 0 if Yes; Not Applicable was not included |
| **In the past month, have you increased, maintained or decreased the frequency of telecommuting (working remotely or working from home)?** | x | | 1 if Decreased; 0 if increased; Did not Change and NA were not included |

**sTable 1. Scoring of COVID related risk behavior and risk perception (cont'd)**

| Question in survey | Behavioral risk | Risk perception | Score |
|---|---|---|---|
| **In the past month, have you increased, maintained or decreased the frequency of handwashing? My frequency of handwashing...** | x | | 1 if Decreased; 0 if increased; Did not Change was not included |
| **In the past month, have you gathered in groups with 10 or more people?** | | | |
| Indoors | x | | 1 if Yes; 0 if No |
| Outdoors | x | | 1 if Yes; 0 if No |
| **When you gathered in groups with 10 or more people, did you practice social distancing? This includes staying 6 feet apart, wearing face coverings and avoiding close interactions.** | x | | 1 if No, 0 if Yes |
| **In the past month, how often have you practiced social distancing (keeping six feet apart) with:** | | | |
| People I don't know (for example, other shoppers in stores, staff and other diners at indoor restaurants, commuters on public transit) | x | | 1 if Never or Sometimes; 0 if Always |
| People I know (friends, family beyond your household, coworkers) | x | | 1 if Never or Sometimes; 0 if Always |
| **In the past month, have you spent time in any of the following places?** | | | |
| A hairdresser, salon or barber | x | | 1 if Yes, 0 if NA |
| The inside of a restaurant or bar | x | | 1 if Yes, 0 if NA |
| A patio or outdoor space at a restaurant or bar | x | | 1 if Yes, 0 if NA |
| An indoor movie theater | x | | 1 if Yes, 0 if NA |
| A shopping mall | x | | 1 if Yes, 0 if NA |
| A church, synagogue, mosque or other place of worship | x | | 1 if Yes, 0 if NA |
| The inside of a house that is not your own | x | | 1 if Yes, 0 if NA |
| A public swimming area such as the pool, lake, ocean or bay | x | | 1 if Yes, 0 if NA |
| A public park | x | | 1 if Yes, 0 if NA |
| A mass gathering like a demonstration or public protest | x | | 1 if Yes, 0 if NA |
| A mass gathering like a political rally | x | | 1 if Yes, 0 if NA |
| A hotel or other short-term rental (like Airbnb) where people outside of your household are staying | x | | 1 if Yes, 0 if NA |
| An overnight trip to another town or city | x | | 1 if Yes, 0 if NA |
| **How worried are you about the new coronavirus overwhelming hospitals?** | | x | 1 if Not at all worried or Not too worried, 0 if Somewhat worried Very worried |
| **How worried are you about getting sick from the new coronavirus again?** | | x | 1 if Not at all worried or Not too worried, 0 if Somewhat worried Very worried |
| **How worried are you about getting sick from the new coronavirus?** | | x | 1 if Not at all worried or Not too worried, 0 if Somewhat worried Very worried |
| **How worried are you about your loved ones getting sick from the new coronavirus?** | | x | 1 if Not at all worried or Not too worried, 0 if Somewhat worried Very worried |
